# Ilioinguinal versus modified Stoppa approach for open reduction and internal fixation of displaced acetabular fractures: A Protocol for Systematic Review and Meta-Analysis

**DOI:** 10.1101/2021.06.22.21259315

**Authors:** Amit Srivastava, Rajesh Kumar Rajnish, Prasoon Kumar, Rehan Ul Haq, Ish Kumar Dhammi

## Abstract

**Background:** The fracture of the acetabulum is one of the most challenging fractures to manage and operate for orthopaedic surgeons. To get a good surgical outcome, anatomical reduction of fractures and reconstruction of the joint is of utmost importance. To achieve a good postoperative outcome an appropriate surgical approach is necessary to achieve an anatomical reduction of fractures and fewer complications.

**Objective:** The current review aims to compare the outcomes of the ilioinguinal versus modified Stoppa approach for open reduction and internal fixation (ORIF) of displaced acetabular fractures by analyzing the evidence from the current literature.

**Methods:** A systematic review of the literature will be conducted in accordance with the PRISMA guidelines. The primary searches will be conducted on the Medline (PubMed), Embase, Scopus, and Cochrane Library databases, using a pre-defined search strategy. The studies of any design in the English language will be included which compared the outcomes of the ilioinguinal and modified Stoppa approach for ORIF of displaced acetabular fractures and reported at least one outcome of interest. Studies that do not compare the outcomes of the ilioinguinal and modified Stoppa approach for the treatment of displaced acetabular fractures in adults (>18 years of age), case reports, conference abstracts, posters, book chapters, review articles, biomechanical studies, technical tips, cadaveric studies, and articles not in the English language will be excluded. Both qualitative and quantitative analyses will be done. Qualitative analysis will be done using appropriate tables and diagrams. Wherever feasible, quantitative analysis to be done with the appropriate software. The risk-of-bias assessment will be done using the MINORS tool for the non-randomized comparative studies, and The Cochrane Collaboration’s risk-of-bias tool will be used for randomized control trials.

## 1. Background

Acetabulum fracture is one of the most challenging intraarticular fractures to manage and operate for orthopaedic surgeons, to get a good surgical outcome, anatomical reduction of fractures and reconstruction of the joint are of utmost importance. [1] To achieve this an appropriate surgical approach is necessary to minimize complications. [2,3]

The ilioinguinal approach is widely used for the fixation of the pelvis and acetabular fractures, it has been found effective in approaching most of the anterior acetabular fracture patterns. [3-5] However, this exposure is a quite extensive approach and poses risk to the neurovascular structures present in proximity, and other soft tissue related complications. [5-7] While the modified Stoppa intrapelvic approach is considered to be less invasive, avoids the middle window of the ilioinguinal approach and gives better exposure of the quadrilateral plate, medial wall of the acetabulum, and sacroiliac joint. This approach requires identification of the corona mortis and its ligation to prevent excessive bleeding, however, there is a risk of the obturator nerve and superior gluteal artery injury. [8-9]

## 2. Need for review

There are several published literature that compared the clinical outcome and complications of the ilioinguinal and modified Stoppa approach however only a few authors have conducted a systematic review or meta-analysis to compare the outcome of the approaches. [10-12] Therefore, the current review aims to perform a systematic review and meta-analysis from current literature to compare the outcomes of the ilioinguinal versus modified Stoppa approach for ORIF of the anterior pelvic ring and acetabular fractures.

## 3. Objectives

### Primary Objectives

i. To compare the primary outcomes like duration of surgery, anatomical reduction quality, total and individual complications for the ilioinguinal and modified Stoppa approach used for the ORIF of displaced acetabular fractures.

### Secondary Objective

i. Additionally, to compare the intraoperative blood loss, and clinical outcomes for both the surgical approaches.

## 4. PICO framework for the study

i. Participants: a human subject with displaced acetabulum fractures
ii. Intervention: Ilioinguinal approach for the open reduction and internal fixation of acetabulum fracture
iii. Control: modified Stoppa approach for the open reduction and internal fixation of acetabulum fracture
iv. Outcomes: The primary outcomes of interest will be the mean duration of surgery, anatomical reduction quality, total and individual complications (vascular injury, nerve injury, infection, and heterotopic ossification). The secondary outcomes of interest will be intraoperative blood loss and clinical outcomes.

## 5. Methods

This systematic review and meta-analysis will be conducted in accordance with the Preferred Reporting Items for Systematic Reviews and Meta-analysis guidelines (PRISMA). [13]

i. *Review Protocol:* A protocol of the review will be formulated in priority in accordance with the PRISMA-P guidelines. (Appendix I)
ii. *Eligibility Criteria:* The studies of any design in the English language will be included that compared the outcomes of the ilioinguinal approach and modified Stoppa approach for the treatment of displaced acetabular fractures and reported at least one outcome of interest. Studies that do not compare the outcomes of the ilioinguinal and modified Stoppa approach for the treatment of acetabular fractures in adults (>18 years of age), case reports, conference abstracts, posters, book chapters, review articles, biomechanical studies, technical tips, cadaveric studies, and articles not in the English language will be excluded.
iii. *Information Sources & Literature search:* A primary literature search will be conducted on the Medline (PubMed), Embase, Scopus, and Cochrane Library databases, using a pre-defined search strategy (Table-1). A manual secondary search of references from the full-text of all included articles and relevant review articles will be conducted. There will be no initial restrictions on the date or language of publication.
iv. *Study Selection:* All the identified articles will be screened through titles and abstracts for eligibility independently by three authors. After initial screening, full texts of all selected articles will be obtained. Eligible articles will be sorted as per the prespecified inclusion and exclusion criteria. The reasons for the exclusion of those articles for which full-text was obtained will be documented. Any discrepancies in the article selection process will be resolved by mutual agreement.
v. *Data Collection & Data Items:* Data will be extracted on pre-formed data collection forms by two authors independently, a third author will cross-check the data for accuracy. Baseline data items will include:
  - Name of the authors and year of publication
  - Number of patients/cases
  - Study design
  - Surgical approach used
  - Number of patients in each group
  - Mean age of the patients
  - Sex of the patients
  - Mean follow up
  - Primary outcomes and
  - Secondary outcomes of interest.
vi. *Outcome Measures:* The following outcome measures will be evaluated however addition and/or modifications will be made if needed:
  - The primary outcomes of interest will be the mean duration of surgery, anatomical reduction quality, total and individual complications (vascular injury, nerve injury, infection, and heterotopic ossification).
  - The secondary outcomes of interest will be intraoperative blood loss and clinical outcomes.
vii. *Data Analysis and Synthesis:* Both qualitative and quantitative data analysis will be performed. For qualitative data analysis, appropriate tables and data visualization diagrams will be used. Quantitative analysis will be performed if ≥ 2 studies included in this review, reported the values of either of the primary or secondary outcomes of interest. To describe the measure of treatment effects, the mean difference will be used for continuous variables, and odds ratio will be used for dichotomous variables. All the results will be expressed along with 95% confidence intervals. Forest plots will be constructed to visualize the results. The statistical heterogeneity will be determined by using the I^2^ test. Reasons for clinical heterogeneity, if any, will be explored. If the heterogeneity was low (I^2^ <75%) fixed-effects model, otherwise the random-effects model (I^2^ >75%) will be used. Meta-analysis will be performed by using Review Manager Software version 5.4. [14]
viii. *Assessment of Risk of Bias:* The risk-of-bias assessment will be done using the MINORS tool for the non-randomized comparative studies [15], and the Cochrane Collaboration’s risk of bias tool [16] will be used for randomized control trials

**Table-1:**
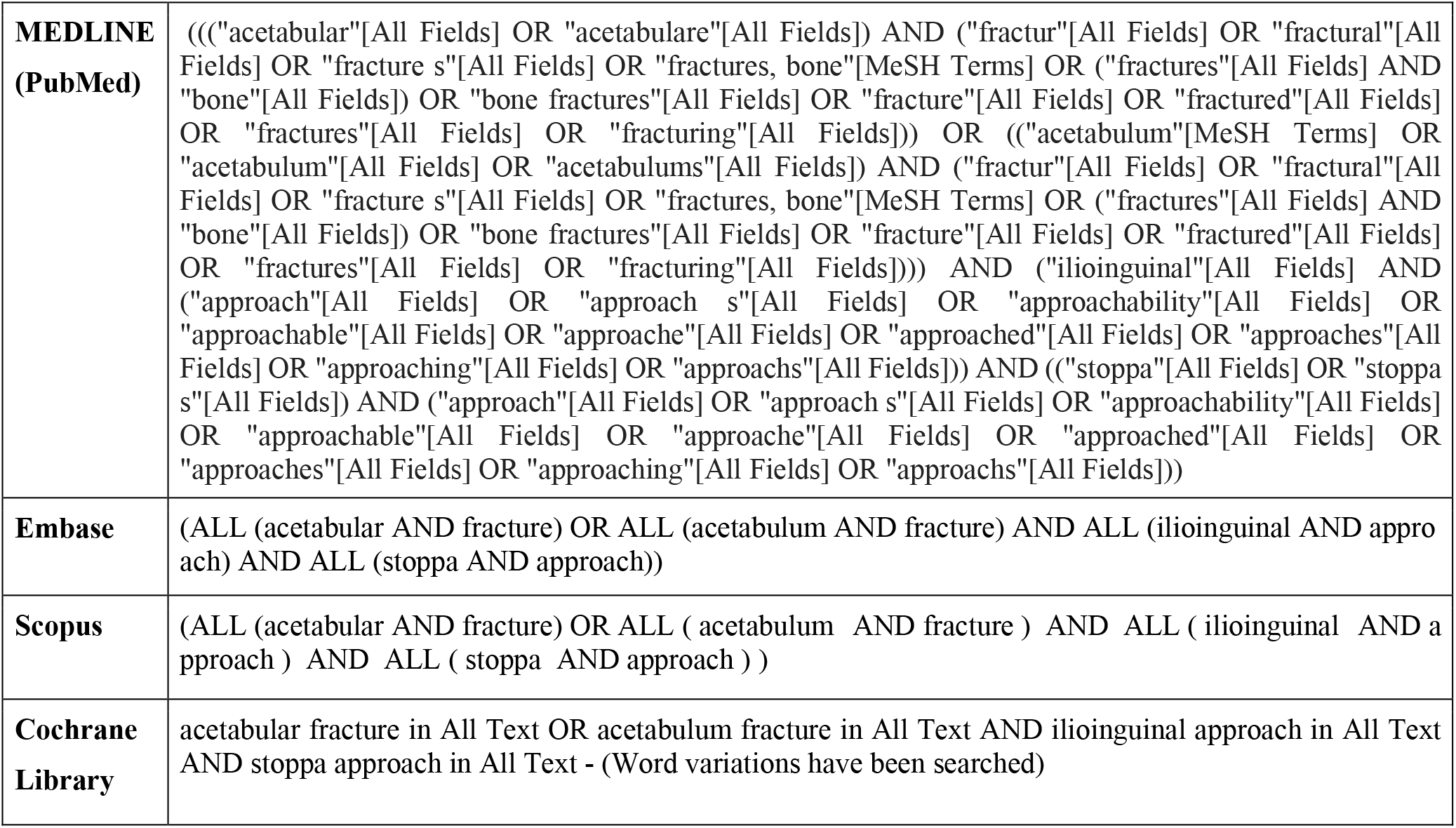
Search Strategy.

## Data Availability

None

## APPENDIX 1: PRISMA-P (Preferred Reporting Items for Systematic review and Meta-Analysis Protocols) 2015 checklist: recommended items to address in a systematic review protocol^*^

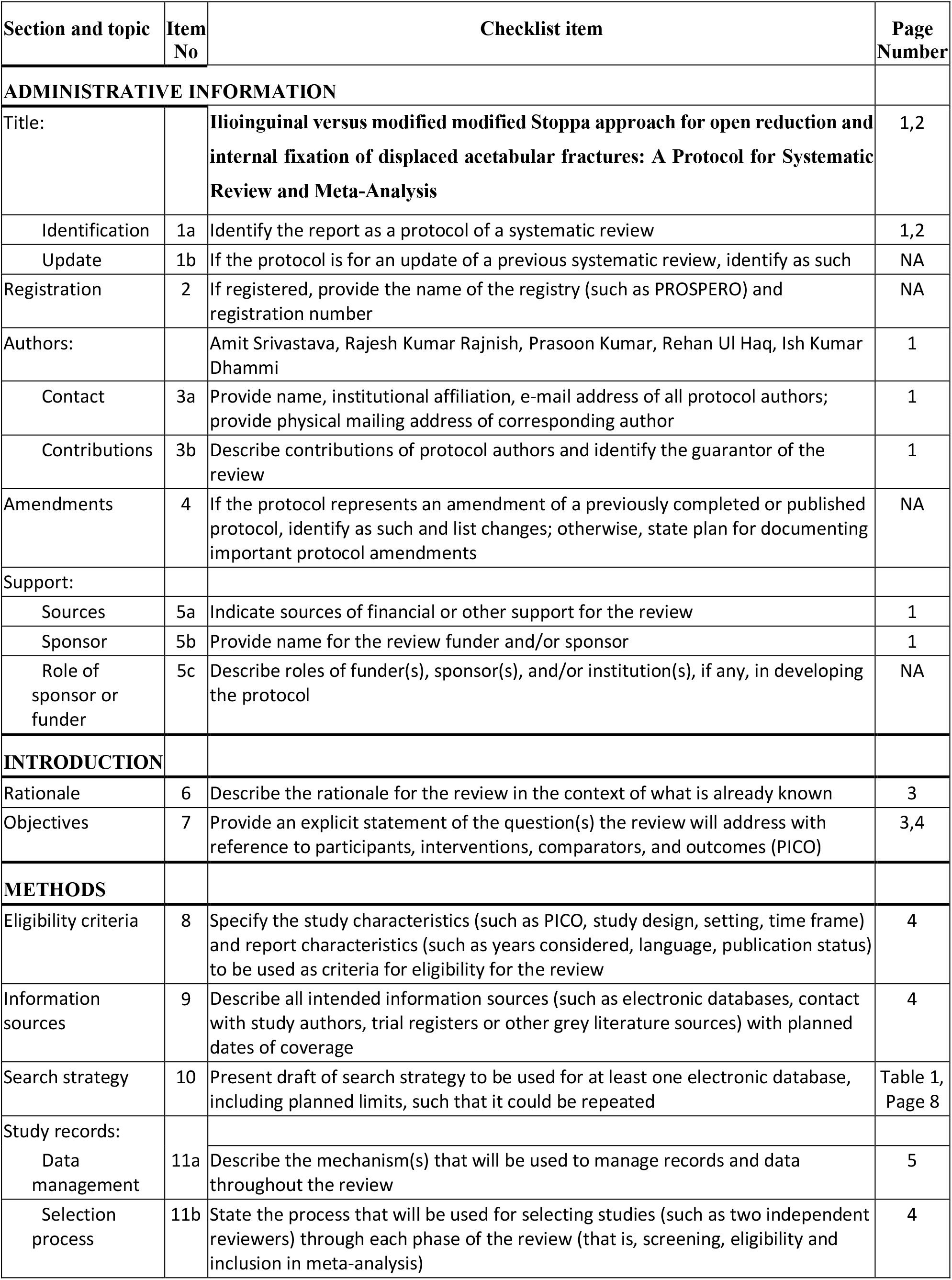
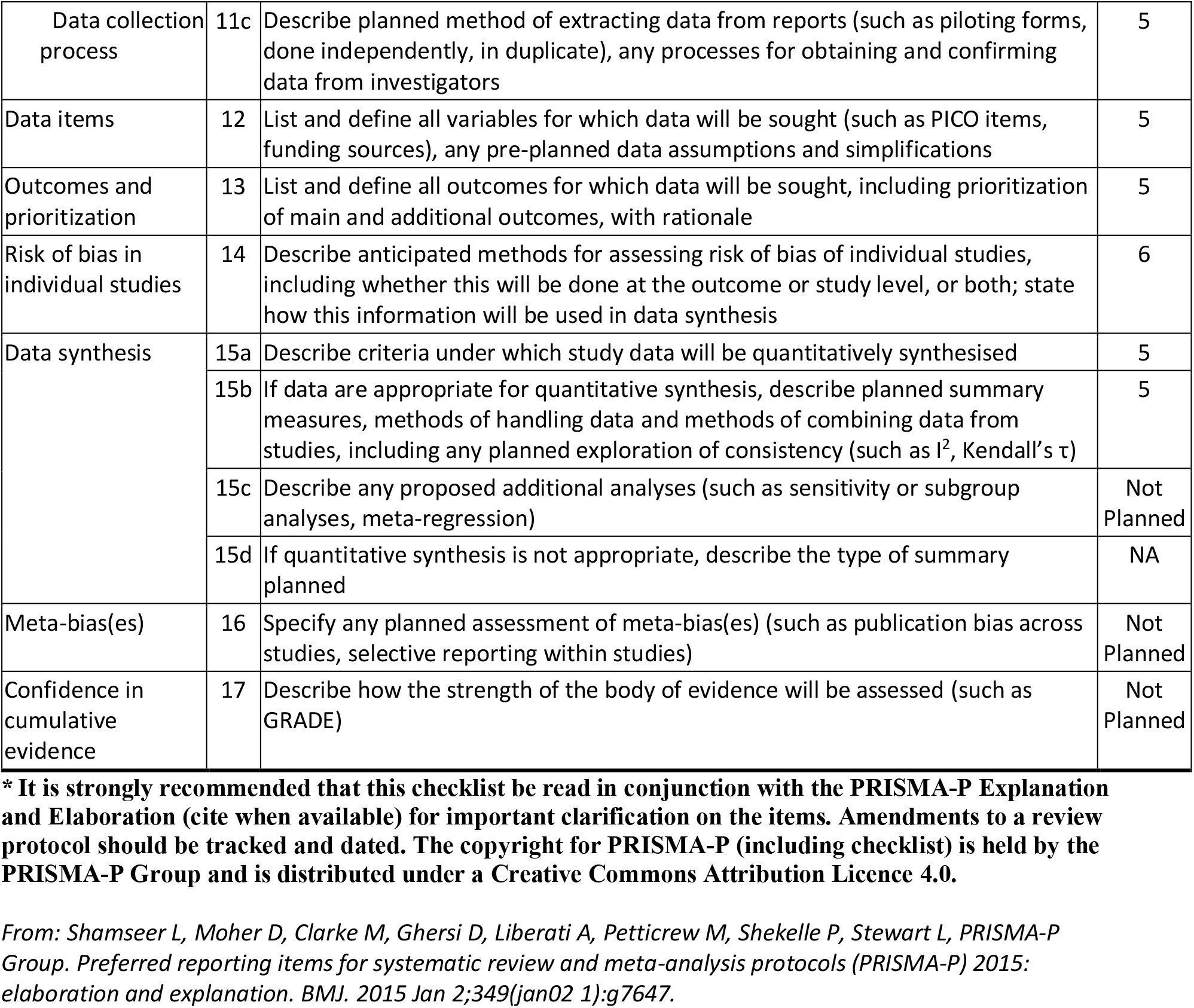

## References

1. Kacra BK, Arazi M, Cicekcibasi AE, Buyukmumcu M, Demirci S. Modified medial Stoppa approach for acetabular fractures: an anatomic study. J Trauma. 2011;71(5):1340–4.

2. Letournel E. The treatment of acetabular fractures through the ilioinguinal approach. Clin Orthop Relat Res. 1993;(292):62–76.

3. Letournel E. Fractures of the acetabulum. A study of a series of 75 cases. 1961. Clin Orthop Relat Res. 1994;(305):5–9.

4. Giannoudis PV, Kanakaris NK, Dimitriou R, Mallina R, Smith RM. The surgical treatment of anterior column and anterior wall acetabular fractures: short-to medium-term outcome. J Bone Joint Surg Br. 2011;93(7):970–4.

5. Seyyed Hosseinzadeh HR, Eajazi A, Hassas Yeganeh M, et al. Modified ilioinguinal approach to the acetabulum and pelvis from beneath the inguinal ligament: a subinguinal approach. Hip Int. 2010;20(2):150–5.

6. Gansslen A, Krettek C. Internal fixation of acetabular both-column fractures via the ilioinguinal approach. Oper Orthop Traumatol. 2009;21(3):270–82.

7. Matta JM. Operative treatment of acetabular fractures through the ilioinguinal approach: a 10-year perspective. J Orthop Trauma.2006;20(1 Suppl): S20–9.

8. Ponsen KJ, Joosse P, Schigt A, Goslings JC, Luitse JS. Internal fracture fixation using the Stoppa approach in pelvic ring and acetabular fractures: technical aspects and operative results. J Trauma. 2006;61(3):662–7.

9. Sagi HC, Afsari A, Dziadosz D. The anterior intra-pelvic (modified rives-stoppa) approach for fixation of acetabular fractures. J Orthop Trauma. 2010;24(5):263–70.

10. Shazar N, Eshed I, Ackshota N, Hershkovich O, Khazanov A, Herman A. Comparison of acetabular fracture reduction quality by the ilioinguinal or the anterior intrapelvic (modified Rives-Stoppa) surgical approaches. J Orthop Trauma. 2014;28(6):313–9.

11. Meena S, Sharma PK, Mittal S, et al. Modified Stoppa approach versus ilioinguinal approach for anterior acetabular fractures; a systematic review and meta-analysis. Bulletin of Emergency and Trauma 2017; 5:6–12.

12. Wu H, Zhang L, Guo X, Jiang X. Meta-analysis of modified Stoppa approach and ilioinguinal approach in anterior pelvic ring and acetabular fractures. Medicine (Baltimore). 2020;99(4):e18395.

13. Page MJ, McKenzie JE, Bossuyt PM, Boutron I, Hoffmann TC, Mulrow CD, et al. The PRISMA 2020 statement: an updated guideline for reporting systematic reviews. BMJ. 2021;372:71.

14. Review Manager (RevMan) [Computer program]. Version 5.4 (2020) The Cochrane Collaboration.

15. Higgins JPT, Altman DG, Gotzsche PC, et al. (2011) Cochrane Bias Methods Group Cochrane Statistical Methods Group. The Cochrane Collaboration’s tool for assessing risk of bias in randomised trials. BMJ 343: d5928.

16. Slim K, Nini E, Forestier D, Kwiatkowski F, Panis Y, Chipponi J. (2003) Methodological index for non-randomized studies (minors): development and validation of a new instrument. ANZ J Surg. 73(9):712–716.

